# Cell-free DNA analysis for the determination of fetal red blood cell antigen genotype in alloimmunized pregnancies

**DOI:** 10.1101/2024.03.26.24304584

**Authors:** Shannon Rego, Olaide ASHIMI Balogun, Kirsten Emanuel, Rachael Overcash, Juan M. Gonzalez, Gregory A. Denomme, Jennifer Hoskovec, Haley King, Ashley Wilson, Julia Wynn, Kenneth J. Moise

## Abstract

**Objective:** The objective of the study is to evaluate the accuracy of NGS based quantitative cfDNA analysis for fetal antigen genotyping in alloimmunized pregnancies undergoing clinical testing in practices across the US as early as ten weeks gestational age. Timely identification of the fetal red blood cell antigen genotype for the antigen to which the pregnant person is alloimmunized is vital for determining risk for hemolytic disease of the fetus or newborn (HDFN) and guiding management. The currently recommended approach to assessing risk for HDFN in the US relies on determining the antigen status of the reproductive partner and/or amniocentesis–an approach with limitations such as low uptake and the potential for nonpaternity, which could be alleviated by the utilization of cfDNA testing to determine fetal antigen status.

**Methods:** Patients with alloimmunized pregnancies undergoing clinical fetal antigen cfDNA analysis were recruited to the study along with the neonates resulting from the pregnancies via outreach from the laboratory. The laboratory issued the results prospectively as a part of clinical care. After delivery, neonatal buccal swabs were sent to an outside, independent laboratory for antigen genotyping. The outside laboratory was blinded to the fetal cfDNA results, and the results were compared. Concordance was reported for the fetal antigen cfDNA analysis for antigens to which the pregnant person was alloimmunized as well as for all antigens for which the pregnant person was genotype negative.

**Results:** A total of 156 participants from 120 clinics who received clinically ordered cfDNA fetal antigen testing provided neonatal buccal swabs for genotyping following delivery. Concordance between cfDNA analysis results and neonatal genotype was determined for 465 antigen calls for the following antigens K1 (n=143), E (124), C (60), Fy^a^ (50), c (47), and D(RhD) (41). These 465 calls included 145 where the fetus was antigen positive and 320 where the fetus was antigen negative. We observed complete concordance between prenatal fetal antigen cfDNA analysis results and neonatal genotypes for the 465 calls, resulting in 100% sensitivity, specificity, and accuracy across a racially and ethnically diverse cohort.

**Conclusion:** This study demonstrates that cfDNA analysis for determining fetal antigen genotype is more accurate than real life application of the current recommendations, i.e., partner testing and amniocentesis, in a diverse US population. In addition, this noninvasive approach reduces barriers to obtaining timely and accurate information about fetal antigen genotype. Taken together with previously published evidence, this study supports the implementation of cfDNA testing to manage alloimmunized pregnancies as a clinically useful and cost effective approach.

## Introduction

Hemolytic disease of the fetus or newborn (HDFN) is a potentially life-threatening form of anemia caused by alloimmunization–a maternal immune response to foreign red blood cell antigens expressed on fetal and neonatal red blood cells inherited from the father.^1^ The American College of Obstetricians and Gynecologists (ACOG) recommends testing the reproductive partner’s antigen status when alloimmunization is diagnosed in pregnancy.^2^ However, rates of reproductive partner screening uptake are low, and results can be complicated by nonpaternity.^3–5^ When reproductive partner antigen status is either unknown or heterozygous, pregnant people are left with the option of amniocentesis for fetal antigen genotyping–an invasive option that carries a risk for fetal loss as well as the chance for an anamnestic increase in the antibody titer or the development of additional alloantibodies. Alternatively, some clinicians may proceed with serial middle cerebral artery peak systolic velocity (MCA-PSV) Doppler measurements, a burdensome screening option that is unnecessary for patients with antigen-negative fetuses.^6^ The need for reproductive partner genotype information also exacerbates an access barrier to testing for populations that already experience disparities in care, as uptake of prenatal testing for reproductive partners is lower in underserved populations.^7^

Cell-free DNA (cfDNA) is already utilized as a standard of care in many European countries for determining fetal antigen status and guiding pregnancy management.^8–10^ In September of 2022, a cfDNA assay utilizing next-generation sequencing (NGS) and quantitative counting template (QCT) technology for determining fetal antigen status was first offered clinically in the US.^11^ The assay improves upon European assays by combining next generation sequencing (NGS) with QCT technology, which facilitates the detection and absolute quantification of variants that are more common in the diverse US population.^11^

In a prior study, we performed initial validation of NGS-based cfDNA analysis with QCT technology for fetal antigen genotyping. While the validation demonstrated 100% sensitivity and specificity of the assay on 1,061 preclinical samples and precision of 99.9% on 1,683 clinical samples, the number of clinical samples with neonatal genotype or serology outcomes was limited to 23 biobank samples from pregnant individuals and 30 pregnancies with prospectively reported results, which showed 100% concordance with ground-truth outcomes.^11^ Herein we build upon that validation with a prospective study comparing fetal antigen cfDNA analysis results completed as part of clinical care for alloimmunized pregnant individuals with antigen genotypes of the resulting neonate tested at an independent laboratory in a total cohort of 156 pregnancies with 465 antigen calls.

## Methods

### Participant Recruitment

Participants were recruited into an IRB-approved fetal antigen patient registry (Western Central IRB protocol # 20225380). Prospective participants were identified via the clinical laboratory’s quality assurance program or by their provider at collaborating clinical sites. Patients and their neonates were eligible for inclusion in the study if: 1) the patient was clinically confirmed to be alloimmunized to at least one of the following antigens: K1 (Kell), Fy^a^ (also known as Fy(a+)), C, c, E, or D(RhD); 2) the patient underwent fetal antigen cfDNA analysis in the US between September 15, 2022 and December 15, 2023 and spoke English or Spanish. Patients were ineligible if their testing was ordered outside of the US. Prospective participants were contacted by a member of the study team or their local clinical team and invited to learn more about the study. Those who agreed to participate provided written informed consent for themselves and their neonate. The participants were compensated for their participation.

### Fetal Antigen cfDNA Assay

Details of the fetal antigen cfDNA analysis have been previously published.^11^ Briefly, we developed and validated an approach to noninvasive prenatal testing that utilizes NGS and QCTs to determine fetal antigen genotypes by analyzing cfDNA in plasma samples from pregnant individuals. The addition of QCTs enables the absolute quantification of detected fetal antigen molecules, which then is compared with the expected number of fetal molecules based on fetal fraction to determine the fetal genotype. The fetal genotype can then be used to predict the fetal antigen phenotype. When the predicted fetal phenotype is antigen positive for an antigen to which the pregnant person is alloimmunized, the pregnancy is at risk for HDFN. This test can be performed as early as ten weeks gestation to determine fetal antigen status in pregnant people who are alloimmunized to the following antigens: K1, Fy^a^, C, c, E, and/or D(RhD). Results are only reported clinically for the antigens to which the patient is alloimmunized. All samples were run on the same version of the fetal antigen cfDNA analysis; the assay did not change during the duration of the study.

### Neonatal genotyping

After receiving informed consent a buccal swab was obtained from the neonate resulting from the alloimmunized pregnancy using ORAcollectDNA buccal swabs (DNA Genotek Inc., Ottawa, Canada). The samples were sent to Grifols Laboratory Solutions Inc (San Marcos, TX), which performed antigen genotyping utilizing BGG Navigator, a polymerase chain reaction (PCR) and genomic hybridization-based genotyping test utilizing ID CORE XTTM™ technology (Progenika Biopharma, S.A., Bizkaia, Spain). Grifols Laboratory was blinded to the fetal cfDNA results. Neonatal genotype and predicted phenotype were reported for the following antigens included in fetal antigen cfDNA analysis: K1, Fy^a^, C, c, and E. For pregnancies alloimmunized to the D(RhD) antigen, neonatal genomic DNA extracted from the swabs was used to amplify exons 1-10 and their flanking regions of the *RHD* gene along with amplification of a hybrid *RHD-CE* exon 3-intron 3 region, and sequenced using BigDye® Terminator v3.1 Cycle Sequencing kit (Applied Biosystems, Foster City, CA) to determine the *RHD* genotype and predicted RhD phenotype.

### Concordance determination

Concordance was determined separately for 1) only those antigens to which the pregnant person was alloimmunized and 2) all antigens for which the pregnant person was genotype negative. A pregnant person must be genotype negative for an antigen (not express the antigen) to be alloimmunized to it. Alloimmunization status does not impact the assay performance as the assay is genotype and not protein (antigen or antibody) based. Therefore, by examining all antigens for which a pregnant person was genotype negative we were able to demonstrate the assay performance with a larger sample size of antigen calls. The researchers and Grifols Laboratory staff were blinded to neonatal and fetal analysis results, respectively, until both assays had been completed. Antigen genotypes were considered concordant where fetal antigen cfDNA analysis predicted fetal phenotype (reported as antigen detected or antigen not detected) and Grifols Laboratory’s neonatal predicted phenotype (reported as antigen positive or antigen negative) were respectively detected and positive OR not detected and negative. If a pregnancy was a twin gestation, the cfDNA analysis results were considered concordant if both neonates were antigen genotype negative and the cfDNA analysis predicted antigen not detected or at least one neonate was antigen genotype positive and the cfDNA analysis result predicted antigen detected.

### Statistics

A sample size of 200 alloimmunized cfDNA assays was selected based on conservative predicted sensitivity of 97% and published antigen allele frequencies to allow for the calculation of the assay analytics with a type I error of up to 5% and a marginal error of 5% (span of 95% confidence intervals). Demographic statistics including maternal age, gestational age, maternal race and ethnicity, and fetal fraction were calculated. As only cases with a cfDNA fetal antigen result were eligible for the concordance study, call rates and turnaround time were calculated for 1,399 consecutive samples sent for fetal antigen cfDNA analysis for the indication of alloimmunization.

## Results

### Fetal Antigen Patient Registry Demographics

The study included 156 participants from 120 different US practices in 37 states who provided written informed consent and submitted neonatal buccal swab samples (see Figure 1, Supplemental Figure 1). The sample included four twin pregnancies. The median gestational age (GA) at the time of testing was 16.4 weeks with a median fetal fraction of 11.1% (Table 1). The pregnancy characteristics and patient demographics, including the breakdown of alloimmunized antigens and antigen detected vs antigen not detected results, were similar to that of a complete patient sample (both enrolled and not enrolled; data not shown). For 1,399 consecutive samples sent for fetal antigen testing the median turnaround time for complete patient samples was 7 days (range 5-21 days), median gestational age was 17 weeks (range 10-39 weeks), and an informative fetal antigen result was returned for 99.6% of cases. Assay call rates were consistent when stratified by patient race and ethnicity and gestational age (data not shown).

**Table 1.** Demographics and pregnancy characteristics of n=156 fetal antigen patient registry participants

| Self-reported race and ethnicity | **n** | **%** |
| --- | --- | --- |
| Asian | 7 | 4.5 |
| Black | 14 | 9.0 |
| Latina/Hispanic | 24 | 15.4 |
| More than one race and/or ethnicity | 2 | 1.3 |
| Unknown | 7 | 4.5 |
| White | 102 | 65.4 |
|  | **Median** | **Range** |
| Maternal age at Estimated Due Date (years) | 31 | 18 - 44 |
| Gestational Age at Time of Fetal Antigen Testing (weeks) | 16.4 | 10.0 - 37.0 |
| Fetal Fraction (%) | 11.1% | 2.4 - 32.2% |
| Trimester of Fetal Antigen Testing | **n** | **%** |
| 1st Trimester | 52 | 33.3 |
| 2nd Trimester | 77 | 49.4 |
| 3rd Trimester | 27 | 17.3 |

### Fetal Antigen Patient Registry Cohort cfDNA analysis Results and Concordance

The 156 patients were alloimmunized to 191 antigens, and concordance calls were made on 190 alloimmunized antigen calls from 155 patients. The most common alloimmunized antigen was E (n=53, 34.0%). Forty-six patients (29.5%) were alloimmunized to K1, 41 (26.3%) to D(RhD), 27 (17.3%) to C, 20 (12.8%) to c, and 4 (2.6%) to Fy^a^. Thirty-four patients were alloimmunized to more than one antigen (Table 2).

**Table 2.** n=155 pregnant individuals alloimmunized to 190 antigens*

| **Antibodies** | **No. of patients tested** | **No. of patients with concordant results** | **% of patients with concordant results** |
| --- | --- | --- | --- |
| E | 39 | 39 | 100% |
| K | 43 | 43 | 100% |
| C, D | 18 | 18 | 100% |
| D | 19 | 19 | 100% |
| c | 10 | 10 | 100% |
| E, c | 9 | 9 | 100% |
| C | 7 | 7 | 100% |
| FyA | 3 | 3 | 100% |
| E, D | 3 | 3 | 100% |
| FyA, E | 1 | 1 | 100% |
| K, c | 1 | 1 | 100% |
| E, K | 1 | 1 | 100% |
| FyA, C, K | 1 | 1 | 100% |

\*One case of the 156 enrolled was excluded from the analysis of alloimmunized antigen calls due to an inconclusive neonatal genotyping result for the D antigen—the antigen to which the patient was alloimmunized (see results section).

Overall, 91 (47.6%) fetal antigen results were reported clinically as “antigen detected”, meaning the fetal antigen genotype predicted an antigen-positive phenotype, and 100 (52.4%) were “antigen not detected”, meaning the fetal antigen genotype predicted an antigen-negative phenotype (Supplemental Table 1).

A concordance call was possible for 190/191 alloimmunized antigens in 155/156 pregnancies. One case had fetal cfDNA results but inconclusive results on neonatal *RHD* sequencing from the outside laboratory, so a concordance call was not possible. Concordance for the 190 fetal antigen cfDNA analysis calls for antigens to which the patients were alloimmunized was 190/190 (100%) (Table 3). The total number of assays is fewer than recommended by the power calculation; however the desired level of statistical power was still achieved with the margins of error less than 5% for all analyses.

**Table 3.** Concordance between fetal antigen cfDNA genotyping results for alloimmunized antigens and neonatal genotyping, n=155 alloimmunized patients, n=190 alloimmunized antigen calls*

|  | **Neonatal**<br>**Antigen Positive** | **Neonatal**<br>**Antigen Negative** | **Total** |
| --- | --- | --- | --- |
| **cfDNA Fetal Antigen Detected** | 90 | 0 | 90 |
| **cfDNA Fetal Antigen Not**<br>**Detected** | 0 | 100 | 100 |
| **Total** | 90 | 100 | 190 |
|  | **%** | **95% Confidence Interval** |  |
| **Sensitivity** | 100% | 96.0% - 100% |  |
| **Specificity** | 100% | 96.4% - 100% |  |
| **PPV** | 100% | 96.0% - 100% |  |
| **NPV** | 100% | 96.4% - 100% |  |
| **Accuracy** | 100% | 98.1% - 100% |  |

\*One case of the 156 enrolled was excluded from the analysis of alloimmunized antigen calls due to an inconclusive neonatal genotyping result for the D antigen—the antigen to which the patient was alloimmunized (see results section).

Additionally, concordance was 100% for 465 fetal antigen calls on antigens to which the pregnant person was genotype negative (Table 4). These 465 calls include 190 calls for antigens to which the pregnant person was alloimmunized, as well as an additional 275 calls on antigens to which the pregnant person was not alloimmunized but for which they were genotype negative (and therefore able to become alloimmunized to the antigen; Table 4, Supplemental Table 2).

**Table 4.**
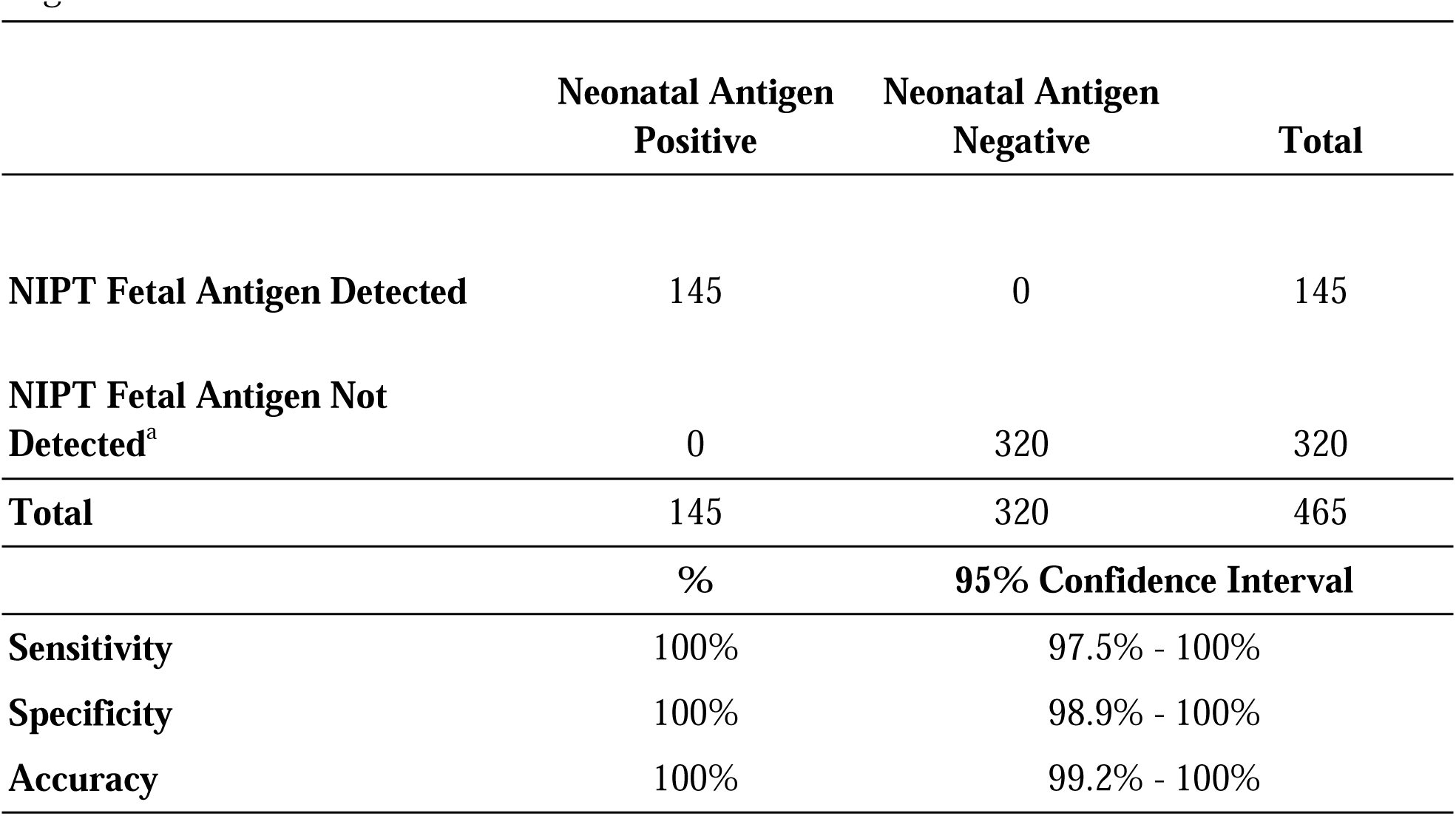
Concordance between fetal antigen NIPT results for all antigens for which the pregnant person is genotype negative in the fetal antigen patient registry cohort, n=156 alloimmunized patients*, n=465 fetal antigen calls for antigens for which the pregnant person was genotype negative.

There was one fetus for which cfDNA analysis correctly called D(RhD) negative due to an *RHD*Ψ variant, which was confirmed via postnatal *RHD* sequencing showing the neonate to be compound heterozygous for the *RHD*-gene deletion and *RHD*Ψ variant.

There were two cases where the cfDNA analysis reported “not detected” for the C antigen and the neonatal genotyping revealed the hybrid RHD-CE-D allele *RHD*DIIIa-ceVS.03(4-7)-RHCE*ce*, present in approximately 0.3% of the US population and associated with an extremely weak C phenotype.^12,13^ Individuals with this weak C phenotype have been documented to produce anti-C antibodies when exposed to C antigen.^12^ Therefore the cfDNA “not detected” call for this genotype is likely an appropriate determination that the fetus is not at risk for HDFN when the pregnant person is alloimmunized to C. However, there is no literature to confirm this, so these cases were excluded from the final calculation of concordance (Table 4). In these cases, the pregnant person was not alloimmunized to C, so no further clinical investigation was warranted.

## Discussion

### Principal findings

This study demonstrates that NGS-based cfDNA analysis using QCT technology is a highly accurate approach for determining fetal antigen genotypes and predicted phenotypes for alloimmunized pregnancies, improving upon the current recommended approach to care in the US. The addition of QCTs enables the absolute quantification of detected fetal antigen molecules. By comparing the detected number of fetal antigen cfDNA molecules with the expected number of molecules based on the fetal fraction, the assay ensures high sensitivity and specificity for the determination of fetal genotype for early gestational ages and low fetal fractions.^11^

In this study concordance between fetal antigen genotype as determined by cfDNA analysis and neonatal antigen genotype as determined by an outside laboratory was 100% for all 190 calls on antigens to which the pregnant person was alloimmunized. Concordance was also 100% when the antigen calls were expanded to include all 465 antigens for which the pregnant person was genotype negative, resulting in an assay sensitivity and specificity of 100%.

The accuracy of fetal antigen cfDNA analysis using NGS with QCT technology is better than the performance of cfDNA analysis utilizing real-time PCR, the approach that has long been used in clinical practice in several countries outside the US for determining fetal antigen status.^9,10,14^ European-based assays have not been adopted in the US due to concerns about performance– both accuracy and inclusivity–for the diverse US population.^15^ The real-time PCR-based assays cannot precisely measure the depth of amplification and therefore cannot quantify the RHD gene molecules. Many of these assays rely on the assumption that the pregnant individual is RhD-negative due to an RHD-gene deletion and therefore, if the control gene indicates the presence of fetal DNA, it is concluded that any RHD gene amplified is of fetal origin, and if no RHD gene is amplified it is concluded that the fetus is RhD-negative. These assumptions can result in false positives or inconclusive results when a non-RHD-gene-deletion genotype is present, as well as false negatives at early gestational ages when fetal fractions are lower.^9,16^ The NGS-based multi-exon sequencing cfDNA analysis for fetal antigen genotyping evaluated in this study uses QCT technology to detect and quantify RHD-negative genotypes including the common RHD-gene deletion as well as variants such as RHDΨ and the RHD-CE hybrid genes which are present in up to 50% of RhD-negative Black Americans.^17^ As a result, this assay has higher call rates than European-based assays. Across over 1,300 clinical samples, an informative fetal antigen result was returned for 99.6% of cases, and call rates were the same across races, ethnicities, and gestational ages (data not shown). An ongoing Phase III trial (clinicaltrials.gov # NCT05912517) for the prevention of HDFN requires enrollment at 13-16 weeks gestation, highlighting the importance of a fetal-antigen cfDNA assay with high performance at early gestational ages (e.g., <15 weeks).^18^ This assay’s unique approach of using absolute quantification enabled by QCTs results in high sensitivity and specificity independent of fetal fraction or gestational age.^11^ Finally, the cfDNA assay simultaneously detects multiple fetal antigens, which is significant as alloimmunization for multiple antigens has implications for worsening fetal anemia.^19^

### Clinical implications

The current recommended approach for determining fetal antigen genotype in alloimmunized pregnancies in the US relies on reproductive partner antigen testing. Amniocentesis is indicated to determine fetal genotype when it cannot be assumed from reproductive partner antigen status.^2^ There are many limitations to this approach in terms of accuracy, feasibility, and utility. Rates of partner testing completion are variable, but in one study, less than a third (12/39) of partners completed the testing.^5^ If reproductive partner testing is not completed, ACOG recommends amniocentesis as a next step. However, amniocentesis carries the risk of fetal loss, worsening alloimmunization due to fetomaternal hemorrhage, and low uptake due to anxiety over the procedure and the risk to the fetus.^20–23^ Subsequently, the fetal antigen status remains unknown for many of these pregnancies; and the pregnancy is then monitored with serial antibody titers and MCA-PSV Doppler ultrasound by specialty providers, a process that is time-intensive and burdensome both for the pregnant person and the healthcare system. These approaches to monitoring have limitations. For example, maternal antibody titers are not useful for monitoring patients who have had prior pregnancies affected with HDFN or who are sensitized to the K antigen because of a lack of correlation between maternal titers and fetal anemia.^2^ In addition, maternal antibody titers are nonspecific and can rise even when the fetus is antigen negative.^24^ MCA-PSV Doppler ultrasound have a sensitivity of 86% and specificity of 71% for pregnancies that have not yet undergone intrauterine transfusions (IUTs) according to a recent meta-analysis by Martinez-Portilla et al, ^25^ and Mari et al. report a false positive rate of 12%.^26^ When MCA-PSV Doppler ultrasound suggests fetal anemia, it can lead to unnecessary and potentially risky invasive procedures such as cordocentesis and IUTs.^6^ While these approaches to monitoring pregnancies at risk for HDFN have limitations, they remain an important and necessary tool for guiding management and assessing the need for intervention when the fetus is antigen positive.

However, cfDNA analysis could mitigate the need for their use in settings where the fetus is not at risk of HDFN due to an antigen-negative genotype, thereby preventing unnecessary cost and logistical challenges of monitoring that is not indicated.

While not identified in this study, the cfDNA assay is designed to maximize sensitivity and therefore there is a small risk for the assay to identify a fetus as antigen positive when the fetus is antigen negative. However, there is no clinical risk to the fetus, and rather this miscall results in unnecessary monitoring; an event that is already occurring with standard care. An additional limitation of the assay is that it does not assess fetal antigen status for all antigens known to cause hemolytic disease, rather, it is designed to determine antigen genotype for those antigens most often implicated in clinically significant HDFN.^1^ The assay also is not designed for use in pregnant people who have had organ or bone marrow transplants or recent blood transfusions.

In addition to the accuracy and logistical burden of the options for managing alloimmunized pregnancies, cost is another important consideration for those making decisions regarding policy and guidelines around the use of cfDNA analysis for determining fetal antigen genotype in alloimmunized pregnancies. To date, only one study has addressed the health economics of cfDNA for the management of alloimmunized pregnancies. The study, by Gajic-Velanoski et al, found a nearly $8,000 ($7,903) reduction in the cost of care when alloimmunized pregnancies were managed using cfDNA fetal genotyping when compared to usual care.^27,28^ The 100% sensitivity and specificity of the NGS-based assay with QCT technology described in this study are higher than those used in Gajic-Veljanoski et al. (99.7% sensitivity and 96.1% specificity), and therefore the cost savings are expected to be even greater with the implementation of this assay.

There was a recent announcement of a US shortage of RhoGAM (a commercially produced Rho(D) immune globulin).^29^ In the US, non-alloimmunized pregnant people who are RhD-negative receive prophylactic Rho(D) immune globulin during pregnancy; however, Rho(D) immune globulin is unnecessary for the approximately 40% of these pregnancies because the fetus is RhD-negative. Multiple European countries currently use cfDNA analysis to determine management for RhD-negative pregnant individuals. Recommended care in the US does not currently include the routine use of cfDNA for this purpose. However, ACOG has noted that it may be an effective and attractive strategy if the inclusivity, feasibility, and cost-effectiveness of cfDNA tests are addressed for the US population and has recently released a practice advisory supportive of the use of cfDNA for RHD fetal antigen genotyping in the context of the Rho(D) immune globulin shortage for the purposes of prioritizing and conserving the supply.^15,30^ The cfDNA analysis described in this study is unimpacted by the alloimmunization status of the pregnancy, and therefore can also be utilized for determining fetal RHD genotype in non-alloimmunized RhD-negative pregnant individuals. This study demonstrates the inclusivity and feasibility of the assay to determine fetal antigen genotype for a diverse US population and supports the use of cfDNA analysis to determine the need for Rho(D) immune globulin to prevent unnecessary administration; conserving a valuable resource for those for whom it is medically necessary.

### Strengths and limitations

This prospective study had sufficient power to demonstrate the accuracy of cfDNA with NGS and QCT technology for fetal antigen genotyping with a high rate of informative results in a diverse US sample of alloimmunized pregnant individuals, including twin pregnancies. The pregnant person’s alloimmunization status was determined through medical records provided by the ordering provider and further confirmed as part of the fetal antigen assay. Importantly, the fetal cfDNA results were reported prospectively as part of the clinical care of the pregnancy without knowledge of the neonatal genotype. The laboratory providing the fetal cfDNA genotyping and the different laboratory performing neonatal genotyping were blinded to each other’s results.

The study enrolled participants from 120 clinical practices across the US including representation from individuals who identified as Asian, Black, Latina, White, and more than one race. Though the study cohort was diverse, participants identifying as White were overrepresented when compared to the US population; however, in contrast to other assays, the no-call rate for this assay did not differ for different races and ethnicities.

While the cost-effectiveness of cfDNA for fetal antigen genotyping has been previously demonstrated^27^, we did not perform an economic analysis as part of this study. Further studies addressing cost-effectiveness in the United States could be beneficial.

### Conclusions

This study demonstrated the accuracy of an NGS-based cfDNA analysis assay with QCT technology for the detection of fetal antigen status in a large, diverse US-based cohort. The performance of this assay is superior to real-life application of the current recommended care in the US. Clinical implementation of fetal antigen cfDNA analysis for the management of alloimmunized pregnancies will remove the barriers of the current recommended care– incomplete partner testing and risk of amniocentesis–resulting in an accurate assessment of fetal risk for more alloimmunized pregnancies, streamlining clinical management, and improving equitable access to care. Taken together with previously published evidence, this study supports the implementation of cfDNA testing to manage alloimmunized pregnancies as a clinically useful and cost-effective approach.

## Conflict of Interest Statement

J.H., J.W., and S.R. are employees of BillionToOne, Inc. and have options/equity in BillionToOne, Inc. H.K. and A.W. are paid contractors with BillionToOne, Inc. K.J.M. is a paid consultant of BillionToOne, Inc. O.A.B., K.E., and R.O. received research funding from BillionToOne, Inc. J.M.G and G.A.D. have no conflicts of interest.

## Data Availability

An anonymized data set is available by request

## Acknowledgements*

We would like to thank the patients who participated in this study. We would also like to thank Emily Griffin, CGC, for her help organizing data; Kelly Chou and John Fang for bioinformatics support; and Oguzhan Atay, Shan Riku, and David Tsao for reviewing the manuscript.

*With the exception of the research participants, all individuals noted in the acknowledgement section are paid employees or contractors with BillionToOne, Inc.

## Author Contributions

Shannon Rego: Methodology, Data Curation, Formal Analysis, Writing - Original Draft, Writing - Review & Editing; Olaide Ashimi Balogun: Investigation, Project Administration; Kirsten Emanuel: Investigation; Rachael Overcash: Investigation, Project Administration; Juan M. Gonzalez: Writing - Review & Editing; Gregory A. Denomme: Investigation, Writing - Review & Editing; Jennifer Hoskovec: Conceptualization, Writing - Review & Editing; Haley King: Investigation, Data Curation; Ashley Wilson: Investigation, Data Curation; Julia Wynn: Conceptualization, Methodology, Writing - Review & Editing, Project Administration; Kenneth J. Moise, Jr: Conceptualization, Methodology, Writing - Review & Editing

## Precis

A cfDNA analysis for fetal antigen genotyping in alloimmunized pregnancies had 100% concordance with neonatal genotype; supporting use of the assay to guide care.

## Authors’ Data Sharing Statement

Anonymized individual participant data will be available. The data will include the pregnant person alloimmunization status, gestational age, fetal fraction, fetal cfDNA results including, if requested, the calibrated fetal antigen fraction (CFAF), and neonate genotype results including the specific genetic variant identified. Data will be available at the time of publication and for 5 years after. Access can be requested by contacting the author, sharing will be determined by the author and the clinical laboratory where the study was conducted. Data will be shared in a secure electronic format for replication purposes.

## Notes

### Competing Interest Statement

J.H., and J.W. are employees of BillionToOne, Inc. and have options/equity in BillionToOne, Inc. H.K. and A.W. are paid contractors with BillionToOne, Inc. K.J.M. is a paid consultant of BillionToOne, Inc. S.R. was an employee of BillionToOne, Inc. O.A.A., K.E., and R.O. received research funding from BillionToOne, Inc. J.M.G and G.A.D. have no conflicts of interest.

### Funding Statement

This study did not receive any funding

### Author Declarations

WCG - IRB a private IRB gave ethical approval for this work. WCG IRB protocol number 20225380

### Summary of Updates

The manuscript has been updated to reflect the changes made following an independent peer review as part of the publication process.

