## Supplemental Tables and Figure for "Cell-free DNA analysis for the determination of fetal red blood cell antigen genotype in alloimmunized pregnancies"

| Table S1: a. cfDNA screening result for alloimmunized antigens, n=190 alloimmunized antigen calls from n=155 pregnancies, b. All cfDNA fetal antigen screening results, n=465 antigen calls from n=156 pregnancies* | | | |
| --- | --- | --- | --- |
| a. |  |  |  |
| **Antigen** | **Detected** | **Not Detected** | **Total** |
| **E** | 19 | 34 | 53 |
| **K** | 5 | 41 | 46 |
| **D** | 31 | 9 | 40 |
| **C** | 15 | 12 | 27 |
| **c** | 17 | 3 | 20 |
| **Fya** | 3 | 1 | 4 |
| **Total** | 90 | 100 | 190 |
| b. |  |  |  |
| **Antigen** | **Detected** | **Not Detected** | **Total** |
| **E** | 34 | 90 | 124 |
| **K** | 5 | 138 | 143 |
| **D** | 31 | 10 | 41 |
| **C** | 27 | 33 | 60 |
| **c** | 33 | 14 | 47 |
| **Fya** | 15 | 35 | 50 |
| **Total** | 145 | 320 | 465 |

*One case of the 156 enrolled was excluded from the analysis of alloimmunized antigen calls due to an inconclusive neonatal genotyping result for the D antigen—the antigen to which the participant was alloimmunized (see results section). It was possible to make concordance calls for that participant on antigens for which she was not alloimmunized but was genotype negative. For this reason there are 155 participants represented in S1a and 156 in S1b.

Table S2: Assay performance by antigen for n=465 concordance calls for antigens to which the pregnant person was genotype negative

| **E antigen** | **n** | **95% Confidence Interval** |
| --- | --- | --- |
| cfDNA antigen detected count | 34 |  |
| cfDNA antigen not detected count | 90 |  |
| Sensitivity | 100% | 89.7% - 100% |
| Specificity | 100% | 96.0% - 100% |
| Accuracy | 100% | 97.1% - 100% |
| **K antigen** | **n** | **95% Confidence Interval** |
| cfDNA antigen detected count | 5 |  |
| cfDNA antigen not detected count | 138 |  |
| Sensitivity | 100% | 47.8% - 100% |
| Specificity | 100% | 97.4% - 100% |
| Accuracy | 100% | 97.5% - 100% |
| **D antigen** | **n** | **95% Confidence Interval** |
| cfDNA antigen detected count | 31 |  |
| cfDNA antigen not detected count | 10 |  |
| Sensitivity | 100% | 88.8% - 100% |
| Specificity | 100% | 69.2% - 100% |
| Accuracy | 100% | 91.4% - 100% |
| **C antigen** | **n** | **95% Confidence Interval** |
| cfDNA antigen detected count | 27 |  |
| cfDNA antigen not detected count | 33 |  |
| Sensitivity | 100% | 87.2% - 100% |
| Specificity | 100% | 89.4% - 100% |
| Accuracy | 100% | 94.0% - 100% |
| **c antigen** | **n** | **95% Confidence Interval** |
| cfDNA antigen detected count | 33 |  |
| cfDNA antigen not detected count | 14 |  |
| Sensitivity | 100% | 89.4% - 100% |
| Specificity | 100% | 76.8% - 100% |
| Accuracy | 100% | 92.5% - 100% |
| **Fya antigen** |  | **95% Confidence Interval** |
| cfDNA antigen detected count | 15 |  |
| cfDNA antigen not detected count | 35 |  |
| Sensitivity | 100% | 78.2% - 100% |
| Specificity | 100% | 90.0% - 100% |
| Accuracy | 100% | 92.9% - 100% |

**Figure S1.** Distribution of participant’s clinics across the United States. Participants were enrolled from 120 different clinics from 37 states. White indicates that no participant was enrolled from a clinic in the state. The dark gray indicates that 4 or more participant clinics were located in that state.

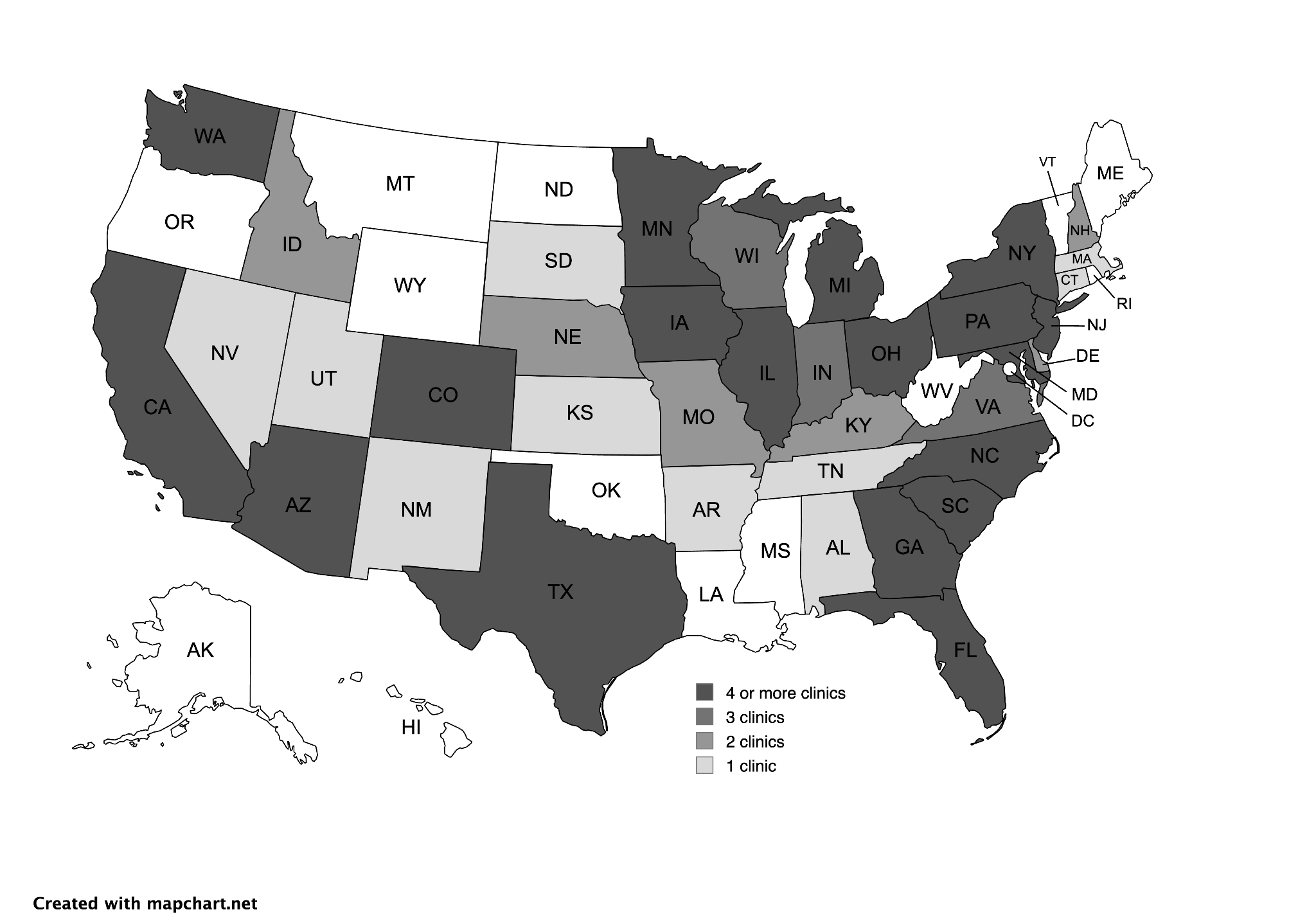
